# Temporal Variations in Seroprevalence of SARS-CoV-2 Infections by Race and Ethnicity in Arkansas

**DOI:** 10.1101/2021.07.15.21260213

**Authors:** Joshua L. Kennedy, J. Craig Forrest, Sean G. Young, Benjamin Amick, Mark Williams, Laura James, Jessica Snowden, Victor M. Cardenas, Danielle Boothe, Catherine Kirkpatrick, Zeel Modi, Katherine Caid, Shana Owens, Marianne Kouassi, Ryan Mann, Claire Putt, Katherine Irish-Clardy, Michael Macechko, Ronald K. Brimberry, Wendy N. Nembhard, Ruofei Du, Jing Jin, Namvar Zohoori, Atul Kothari, Hoda Hagrass, Ericka Olgaard, Karl W. Boehme

## Abstract

**Objective:** Our objective is to estimate CoV-2 infection rates in a rural state using seroprevalence of antibodies to CoV-2 as an indicator of infection.

**Study Design and Setting:** This is a single-site study within an academic center and regional programs within the state of Arkansas. We obtained residual serum samples from a convenience sample of adults who were outpatients and came to the hospital or regional clinic for non-COVID-related reasons. We collected remnant in three time periods (August 15 to September 5, September 12 to October 24, and November 7 to December 19).

**Results:** In 2020, the overall age, gender, and race standardized prevalence of CoV-2 antibodies was 2.6% (August to September), 4.1% (September to October), and 7.4% (November to December). There was no difference in seroprevalence between urban compared to rural areas. Positive tests were not uniformly distributed across racial and ethnic minorities. Higher seroprevalence rates were found in Hispanics and Blacks or African Americans compared to whites across all time periods.

**Conclusions:** In a state with a large rural population, 2.6-7.4% of people experienced CoV-2 infection by December 2020. Blacks and Hispanics had disproportionately higher rates of CoV-2 infections than whites.

**What is new?:** *Key findings:* In this prospective convenience sampling of remnant sera, we found increasing seroprevalence from 2.6% to 7.4% (August 2020 to December 2020). Higher seroprevalence rates were found in Hispanics and Blacks or African Americans compared to whites across all time periods, and no difference was determined between those individuals from rural or urban areas.

*What this adds to what is known:* In a largely rural population, Blacks and Hispanics had disproportionately higher rates of CoV-2 infections than whites, and these populations need to be studied further regarding outcomes.

*What is the implication?:* There are health disparities that exist regarding CoV-2 infections, and we should target vaccination information and education to these groups.

**Highlights:**

- SARS-CoV-2 infections increased from 2.6% to 7.4% from August to December 2020.
- Higher seroprevalence was found in Hispanics and Blacks as compared to whites.
- There was no difference in the seroprevalence in rural compared to urban areas.

## INTRODUCTION

Since emerging in 2019, Severe Acute Respiratory Syndrome Coronavirus-2 (CoV-2) has spread around the world causing high morbidity and mortality[1-4]. CoV-2 infections in the United States (US) were initially concentrated in densely populated areas, but later spread to rural areas[5-7]. Further, significant racial and ethnic disparities in the rates of CoV-2 infection are noted with a higher prevalence in Black and Hispanic persons[4, 8].

With limited testing and inconsistent symptom severity, including a spectrum of asymptomatic to fatal infections, the true prevalence of CoV-2 infections has been difficult to determine. Some reports estimate that 40 to 45% of cases of CoV-2 may be asymptomatic[9, 10]. Asymptomatic cases affect our ability to contain CoV-2 within the population because the virus can be spread surreptitiously to others[11, 12]. Many in the scientific community now recognize the best way to evaluate the effect of asymptomatic cases on CoV-2 spread and to determine when herd immunity may be reached is population-based seroprevalence surveys[13, 14]. These surveys provide valuable information to public health officials as they consider decisions related to the pandemic.

Arkansas, a small state in the southern US with an ethnically- and racially-diverse population of approximately 3.2 million, provides an opportunity to determine CoV-2 seroprevalence in urban vs. rural populations and evaluate racial and ethnic disparities in a temporal fashion. According to US census reports, the four largest ethnic groups in the state are White (Non-Hispanic) 72.1%, Black (Non-Hispanic) 15.1%, White (Hispanic) 4.38%, and Other (Hispanic) 2.66%[9]. This report describes the variations of CoV-2 infection over time (August to December 2020) by age, gender, race/ethnicity, and rurality based on serologic testing in the state.

## METHODS

### Human specimens

Remnants from serum samples collected for routine, non-COVID-19-related outpatient clinical laboratory tests that would otherwise be discarded were obtained from the University of Arkansas for Medical Sciences (UAMS) main campus in central Arkansas and three regional campuses located in northwest (Springdale and Fort Smith) and southeast Arkansas (Pine Bluff). The study was reviewed and approved by the UAMS Institutional Review Board and waiver of consent and HIPAA applied.

Inclusion criteria for serum collection were age 18 years or older, Arkansas resident, and outpatient source. Samples were excluded with the following diagnosis codes: immunodeficiency (primary immune deficiency (D80-D89), transplant recipient (all codes beginning with Z94), and cancer (C00-D49)). In addition, samples from patients with chemotherapy (prior two months), steroids (prior 30 days), and/or intravenous immunoglobulin (prior 6 months) were excluded.

Samples were collected across three time periods. Time period 1 included samples from August 15 to September 5, time period 2 from September 12 to October 24, and time period 3 from November 7 to December 19. For time period 1, samples were collected from UAMS, Pine Bluff, and Springdale (Northwest Arkansas, NWA) sites. Time period 2 and 3 included samples from UAMS, Pine Bluff, Springdale, and Fort Smith. The Arkansas Department of Health also contributed samples from across the state for time period 3.

Criteria for selection of samples were provided to an honest broker to create lists of subjects twice weekly from the UAMS electronic health record, known as the Arkansas Central Data Repository (AR-CDR). Samples were selected from Fort Smith, Pine Bluff, and Northwest Arkansas by staff at each site after they were considered remnant. All clinical and demographic variables were stored in a protected REDCap database[10, 11] and included patient age, gender, race, ethnicity, ZIP Code, and county of residence. Patient ZIP Codes were cross referenced with the Federal Office of Rural Health Policy’s data files identifying non-metropolitan counties and rural census tracts that comprise the rural area in our study[12].

### Laboratory Methods

A two-step process was used to evaluate the sera for CoV-2 antibodies, as suggested by Centers for Disease Control (CDC) guidelines[13]. All sera were tested for CoV-2 Receptor Binding Domain (RBD) IgG antibodies using the Beckman Coulter (BC) Access SARS-CoV-2 IgG (Brea, CA) in the Clinical Laboratory at the UAMS main campus. This immunoassay utilizes paramagnetic beads labeled with recombinant protein for the SARS-CoV-2 RBD of the S1 protein. Bound CoV-2 antibodies are labeled with an alkaline phosphatase conjugate and a reaction with a chemiluminescent material generates light measured by a luminometer. The result is based on the signal to established cut-off point of the assay. The Access SARS-CoV-2 IgG assay has a positive percent agreement (PPA) of 100% >18 days after a positive CoV-2 PCR test and a 99.6% negative percent agreement (NPA) in samples tested prior to COVID-19. Based on studies submitted in their EUA application, Beckman Coulter shows the PPA from 0-7 days after the time of a positive CoV-2 PCR test is 75.8%, from 8-14 days the PPA is 95.3%, and from 15 days or greater is 96.8%. The UAMS Clinical Laboratory conducted an independent verification study and found 74%, 94% and 100%, PPA based on days 0-7, 8-14, and ≥15, respectively, and 100% NPA.

Specimens scored as reactive by BC and 5% of the negative-scoring samples were confirmed using a laboratory-developed enzyme-linked immunosorbent assay (ELISA). A final result of “positive” required that both assays indicated reactivity.

### Four-Antigen Confirmation Test (FACT)

The laboratory-derived assay in this study was designed considering prior ELISAs for CoV-2 antibodies[14]. Plates were coated with CoV-2 RBD, Spike, Nucleoprotein, and bovine serum albumin (BSA) (negative control) diluted to 2μg/mL. Plates were then washed three times with PBS + 0.1% Tween-20 (PBS-T) and blocked for up to 4h with 200 μL of PBS-T + 3% milk. Serum samples were diluted 1:50 in PBS-T + 1% milk. CoV2-positive serum and pre-COVID-19 sera were utilized as a positive and negative control, respectively, and all samples were run in duplicate. Blocking reagent was removed and 50 µL of diluted serum or controls were added in the plates were incubated at RT for 2h. The plates were washed three times with PBS-T and incubated with 50 µL of secondary antibody solution anti-human IgG + IgM (Jackson ImmunoResearch) diluted 1:5000 in PBS-T + 1% milk) for 1h at RT. Then, the plates were washed three times with PBS-T and 75 µL of SureBlue TMB 1-Component Peroxidase Substrate (SeraCare) was added to each well. After 5 min incubation, 75 µL of TMB Stop Solution (SeraCare) was added. The OD450 was measured using a FluoStar Omega plate reader (BMG Labtech). Final RBD OD450 was calculated by subtracting the mean absorbance of BSA from the mean OD450 of RBD duplicate samples. Positives on the assay required at least 2 positive antigens that reached a threshold of 0.3 optical density (OD) after subtracting the BSA result. Pre-COVID-19 sera and sera from PCR-confirmed CoV-2 patients were utilized as positive and negative controls. Sensitivity and specificity based on these samples was 100% and 94.6%, respectively.

### Statistical Analyses

The prevalence of CoV-2 antibodies in the sample was reported with 95% confidence intervals (CI) obtained using exact binomial distributions. Age, gender, and race/ethnicity standardized prevalence rates were calculated using 2019 U.S. Census Bureau Arkansas state adult population estimates. Univariable and multivariable logistic regressions were employed to study associations between variables and the rate of positive antibodies to CoV-2 for each time period separately.

The significance of a monotone trend of the positivity to CoV-2 was tested by the Cochran-Armitage method. A multivariable logistic regression using the backward selection algorithm was employed to study the trend effect of race ethnicity by the time period, starting with main effects and two-way interaction terms, and in each step retaining only the factors showing significant associations (i.e. p < 0.2). The final model includes gender, race/ethnicity, sample collection site, time period, and the two-way interaction terms: time period with race/ethnicity, time period with the site.

Due to the outcome rate <10%, a bias-reducing penalized likelihood optimization was applied for all the logistic regression fittings[15]. Goodness-of-fit was examined by the Deviance test results which did not suggest model fitting concerns. Statistical significance was set at 0.05. All analyses were conducted using SAS, version 9.4. and graphics were done using R, version 4.0.2, and ArcGIS Pro, version 2.7.3.

## RESULTS

### Cohort characteristics

We collected 1,301 adult remnant serum samples from four sites between time period 1, 2098 for time period 2, and 2405 for time period 3 for a total sample collection of 5804. Table 1 provides summary data for demographic characteristics for each time period. While a majority of the samples came from northwest, southeast, and central Arkansas, we did have samples representing 74/75 counties in Arkansas during our collection time points (Figure 1).

**Table 1.** Demographics of patients with CoV-2 antibody test results by time period.

| Characteristics <sup>a</sup> | Time Period 1<br>(N=1301) | Time Period 2<br>(N=2098) | Time Period 3<br>(N=2405) |
| --- | --- | --- | --- |
| Positive cases, n (%) | 49 (3.77) | 103 (4.91) | 194 (8.07) |
| Age group, years, n (%) |  |  |  |
| 18-29 | 257 (19.8) | 406 (19.4) | 562 (23.4) |
| 30-39 | 244 (18.8) | 394 (18.8) | 428 (17.8) |
| 40-49 | 205 (15.8) | 294 (14.0) | 335 (13.9) |
| 50-59 | 211 (16.2) | 335 (16.0) | 358 (14.9) |
| 60-69 | 209 (16.1) | 349 (16.6) | 377 (15.7) |
| 70+ | 175 (13.5) | 319 (15.2) | 345 (14.4) |
| Missing (n) | 0 | 1 | 0 |
| Age, years, mean (SD) | 47.80 (18.17) | 48.60 (18.53) | 47.21 (18.78) |
| Missing (n) | 0 | 19 | 0 |
| Gender, n (%) |  |  |  |
| Female | 898 (69.3) | 1430 (68.2) | 1661 (69.1) |
| Male | 398 (30.7) | 668 (31.8) | 742 (30.9) |
| Missing (n) | 5 | 0 | 2 |
| Race/Ethnicity, n (%) |  |  |  |
| White | 565 (45.7) | 937 (47.6) | 1120 (47.0) |
| Black | 558 (45.2) | 831 (42.2) | 1026 (43.1) |
| Hispanic | 74 (6.0) | 126 (6.4) | 137 (5.8) |
| Others | 39 (3.2) | 75 (3.8) | 99 (4.2) |
| Missing (n) | 65 | 129 | 23 |
| Area, n (%) |  |  |  |
| Urban | 1126 (86.8) | 1838 (87.6) | 1990 (82.8) |
| Rural | 171 (13.2) | 260 (12.4) | 413 (17.2) |
| Missing (n) | 4 | 0 | 2 |
| Site, n (%) |  |  |  |
| UAMS | 981 (75.4) | 1338 (63.8) | 1485 (61.8) |
| Pine Bluff | 266 (20.5) | 403 (19.2) | 319 (13.3) |
| Ft. Smith | - | 206 (9.8) | 131 (5.5) |
| Springdale | 54 (4.2) | 151 (7.2) | 220 (9.2) |
| ADH | - | - | 250 (10.4) |
| COVID-19 test history, n (%) |  |  |  |
| Yes | 139 (11.4) | 271 (12.9) | 437 (20.3) |
| No | 1078 (88.6) | 1827 (87.1) | 1717 (79.7) |
| Missing (n) | 84 | 0 | 251 |
| COVID-19 PCR result, n (%) |  |  |  |
| Positive | 12 (9.1) | 17 (6.3) | 8 (8.3) |
| Negative | 120 (90.9) | 253 (93.7) | 89 (91.8) |
| Missing (n) | 1169 | 1828 | 2308 |

<sup>a</sup> Missing category is not included in the descriptive calculation of % provided in this table.

**Figure 1.**
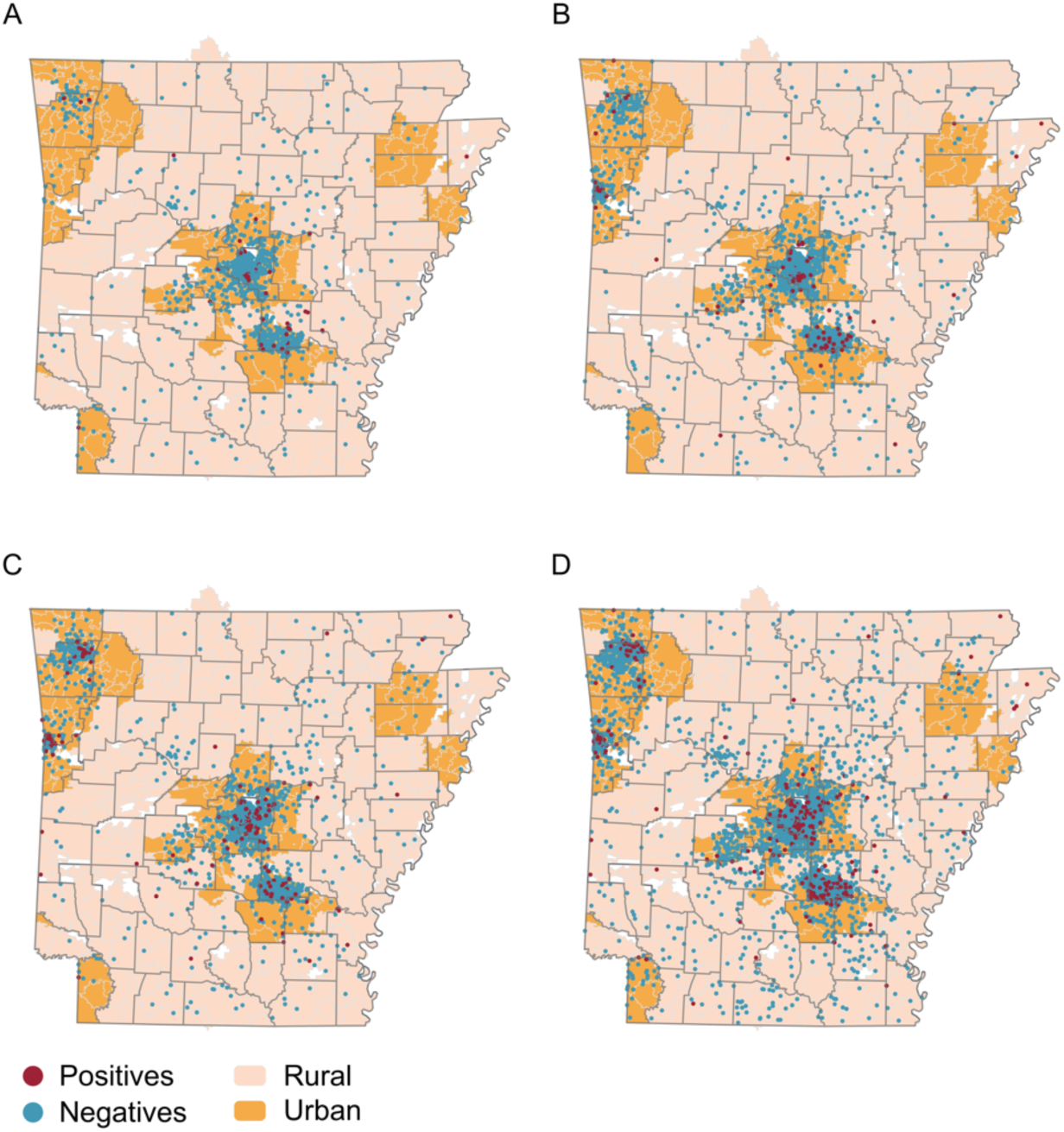
Figure 1 Geographic distribution of study participants. The residences of study enrollees are indicated for (A) Time period 1, (B) Time period 2, (C) Time period 3, and (D) all time periods. Areas considered to be urban are indicated with orange highlight. Red dots and blue dots indicate specimens that tested positive or negative for SARS-CoV-2 antibodies, respectively.

### Overall seroprevalence results in each time period

The observed raw seroprevalence rates were 3.8%, 4.9%, and 8.1% for the three time periods, respectively (Table 2). Both the unadjusted and adjusted association study results show that race/ethnicity is significantly associated with the positive CoV-2 antibodies (Tables 2 and 3). The estimates obtained from the remnant samples were standardized by the distribution of age, gender and race/ethnicity of the Arkansas general population. The standardized rate per 100 persons was 2.6% (95%CI 1.7-3.5), for time period 1. For time period 2, we found a standardized seroprevalence of 4.1% (95%CI 3.1-5.1). Seroprevalence increased slightly in time period 3 where we saw an age-, gender, and race/ethnicity-standardized rate of 7.4% (95%CI 6.0-8.7) (Figure 2).

**Table 2.** Unadjusted association to positivity of CoV-2 antibody examined by univariable logistic regression at each time period.

|  | Time Period 1 (N=1301) |  |  | Time Period 2 (N=2098) |  |  | Time Period 3 (N=2405) |  |  |
| --- | --- | --- | --- | --- | --- | --- | --- | --- | --- |
|  | Positive cases N (%) | OR (95%CI) | P-value<sup>c</sup> | Positive cases N (%) | OR (95% CI) | p-value<sup>c</sup> | Positive cases N (%) | OR (95% CI) | P-value<sup>c</sup> |
| Total, N (%)<sup>a</sup> | 49 (3.8) |  |  | 103 (4.9) |  |  | 194 (8.1) |  |  |
| Age, N (%) |  |  | 0.0526 |  |  | 0.1664 |  |  | 0.4081 |
| 18-29 | 17 (6.6) | 3.59 (1.12, 11.52) |  | 22 (5.4) | 1.57 (0.76, 3.25) |  | 50 (8.9) | 1.65 (0.96, 2.84) |  |
| 30-39 | 13 (5.3) | 2.87 (0.87, 9.50) |  | 29 (7.4) | 2.17 (1.08, 4.36) |  | 40 (9.4) | 1.75 (1.00, 3.06) |  |
| 40-49 | 6 (2.9) | 1.61 (0.43, 6.00) |  | 13 (4.4) | 1.29 (0.58, 2.88) |  | 24 (7.2) | 1.32 (0.71, 2.44) |  |
| 50-59 | 7 (3.3) | 1.81 (0.50, 6.56) |  | 14 (4.2) | 1.21 (0.55, 2.67) |  | 32 (8.9) | 1.67 (0.93, 2.99) |  |
| 60-69 | 3 (1.4) | 0.84 (0.19, 3.74) |  | 13 (3.7) | 1.08 (0.48, 2.40) |  | 29 (7.7) | 1.42 (0.78, 2.56) |  |
| 70+ | 3 (1.7) | Ref |  | 11 (3.5) | Ref |  | 19 (5.5) | Ref |  |
| Gender, N (%) |  |  | 0.1325 |  |  | 0.7249 |  |  | 0.1827 |
| Female | 39 (4.3) | 1.70 (0.85, 3.40) |  | 72 (5.0) | 1.08 (0.70, 1.66) |  | 126 (7.6) | 0.81 (0.60, 1.10) |  |
| Male | 10 (2.5) | Ref |  | 31 (4.6) | Ref |  | 68 (9.2) | Ref |  |
| Race/Ethnicity, N (%) |  |  | <.0001 |  |  | <.0001 |  |  | <.0001 |
| White | 6 (1.1) | Ref |  | 24 (2.6) | Ref |  | 62 (5.5) | Ref |  |
| Black | 27 (4.8) | 4.45 (1.88, 10.56) |  | 45 (5.4) | 2.16 (1.31, 3.56) |  | 91 (8.9) | 1.66 (1.19, 2.31) |  |
| Hispanic | 13 (17.6) | 18.89 (7.13, 50.09) |  | 26 (20.6) | 9.83 (5.46, 17.70) |  | 32 (23.4) | 5.22 (3.26, 8.35) |  |
| Others | 0 (0.0) | 1.09 (0.06, 20.39) |  | 5 (6.7) | 2.91 (1.11, 7.61) |  | 7 (7.1) | 1.37 (0.62, 3.03) |  |
| Area, N (%) |  |  | 0.6351 |  |  | 0.6646 |  |  | 0.1713 |
| Urban | 44 (3.9) | 1.24 (0.50, 3.07) |  | 92 (5.0) | 1.15 (0.61, 2.15) |  | 154 (7.7) | 0.776 (0.54, 1.12) |  |
| Rural | 5 (2.9) | Ref |  | 11 (4.2) | Ref |  | 40 (9.7) | Ref |  |
| Site, N (%) |  |  | 0.5530 |  |  | 0.7787 |  |  | <.0001 |
| UAMS | 35 (3.6) | Ref |  | 63 (4.7) | Ref |  | 97 (6.5) | Ref |  |
| Pine Bluff | 11 (4.1) | 1.20 (0.61, 2.37) |  | 22 (5.5) | 1.19 (0.72, 1.94) |  | 18 (5.6) | 0.87 (0.52, 1.46) |  |
| Ft. Smith | - | - |  | 12 (5.8) | 1.29 (0.69, 2.42) |  | 26 (19.9) | 3.58 (2.23, 5.75) |  |
| Springdale | 3 (5.6) | 1.81 (0.58, 5.68) |  | 6 (4.0) | 0.90 (0.39, 2.05) |  | 20 (9.1) | 1.46 (0.88, 2.40) |  |
| ADH | - | - |  | - | - |  | 33 (13.2) | 2.19 (1.44, 3.33) |  |
| COVID-19 test history, N (%) |  |  | 0.0007 |  |  | 0.2293 |  |  | 0.0528 |
| Yes | 12 (8.6) | 3.26 (1.65, 6.45) |  | 17 (6.3) | 1.39 (0.81, 2.36) |  | 42 (9.6) | 1.44 (0.996, 2.08) |  |
| No | 31 (2.9) | Ref |  | 86 (4.7) | Ref |  | 119 (6.9) | Ref |  |
| COVID-19 PCR result, N (%) |  |  | <.0001 |  |  | <.0001 |  |  | 0.0307 |
| Positive | 10 (83.3) | 199.06 (29.52, >999.99) |  | 14 (82.4) | 296.46 (59.86, >999.99) |  | 4 (50.0) | 5.21 (1.17, 23.26) |  |
| Negative | 2 (1.7) | Ref |  | 3 (1.2) | Ref |  | 14 (15.7) | Ref |  |

<sup>a</sup> missing data are NOT included in the analysis for a variable.
<sup>b</sup> % in 'Final COVID-19 test result' is by Positive/Negative for a category of a variable, i.e. sum of the row % for a category equals to one.
<sup>c</sup> *p*-value is for the testing whether the positivity of Final test result differs between the category of a respective variable.

**Table 3.** Adjusted association to positivity of CoV-2 antibody examined by multivariable logistic regression at each time period.

| Effect | Time Period 1 (N=1301) |  | Time Period 2 (N=2098) |  | Time Period 3 (N=2405) |  |
| --- | --- | --- | --- | --- | --- | --- |
|  | OR (95%CI) | P-value | OR (95%CI) | P-value | OR (95%CI) | P-value |
| Age |  | 0.4392 |  | 0.6390 |  | 0.8960 |
| 18-29 vs 70+ | 2.04 (0.62, 6.65) |  | 1.09 (0.51, 2.3) |  | 1.14 (0.64, 2.04) |  |
| 30-39 vs 70+ | 1.84 (0.55, 6.14) |  | 1.66 (0.8, 3.42) |  | 1.29 (0.71, 2.31) |  |
| 40-49 vs 70+ | 1.08 (0.28, 4.16) |  | 1.13 (0.51, 2.54) |  | 0.95 (0.51, 1.79) |  |
| 50-59 vs 70+ | 1.4 (0.38, 5.14) |  | 1.17 (0.53, 2.58) |  | 1.19 (0.65, 2.17) |  |
| 60-69 vs 70+ | 0.71 (0.16, 3.09) |  | 1.01 (0.45, 2.26) |  | 1.21 (0.67, 2.21) |  |
| Gender |  | 0.3658 |  | 0.9875 |  | 0.1131 |
| Female vs Male | 1.39 (0.68, 2.83) |  | 1 (0.63, 1.57) |  | 0.77 (0.56, 1.06) |  |
| Race/Ethnicity |  | <.0001 |  | <.0001 |  | <.0001 |
| Black vs White | 3.9 (1.66, 9.19) |  | 1.93 (1.14, 3.26) |  | 2.03 (1.41, 2.92) |  |
| Hispanic vs White | 15.06 (5.59, 40.6) |  | 9.83 (5.32, 18.14) |  | 4.42 (2.71, 7.23) |  |
| Others vs White | 0.91 (0.05, 15.63) |  | 2.55 (0.98, 6.65) |  | 1.43 (0.64, 3.17) |  |
| Area |  | 0.5473 |  | 0.8258 |  | 0.1481 |
| Urban vs Rural | 0.75 (0.3, 1.9) |  | 0.93 (0.47, 1.81) |  | 0.73 (0.47, 1.12) |  |
| Site |  | 0.5701 |  | 0.4906 |  | <.0001 |
| Pine Bluff vs UAMS | 1.49 (0.71, 3.14) |  | 1.49 (0.87, 2.54) |  | 0.8 (0.47, 1.37) |  |
| Ft. Smith vs UAMS | - |  | 0.85 (0.41, 1.79) |  | 3.37 (1.99, 5.71) |  |
| Springdale vs UAMS | 0.89 (0.21, 3.78) |  | 1.01 (0.42, 2.43) |  | 1.73 (1.01, 2.97) |  |
| ADH vs UAMS | - |  | - |  | 1.75 (1.07, 2.85) |  |

**Figure 2.**
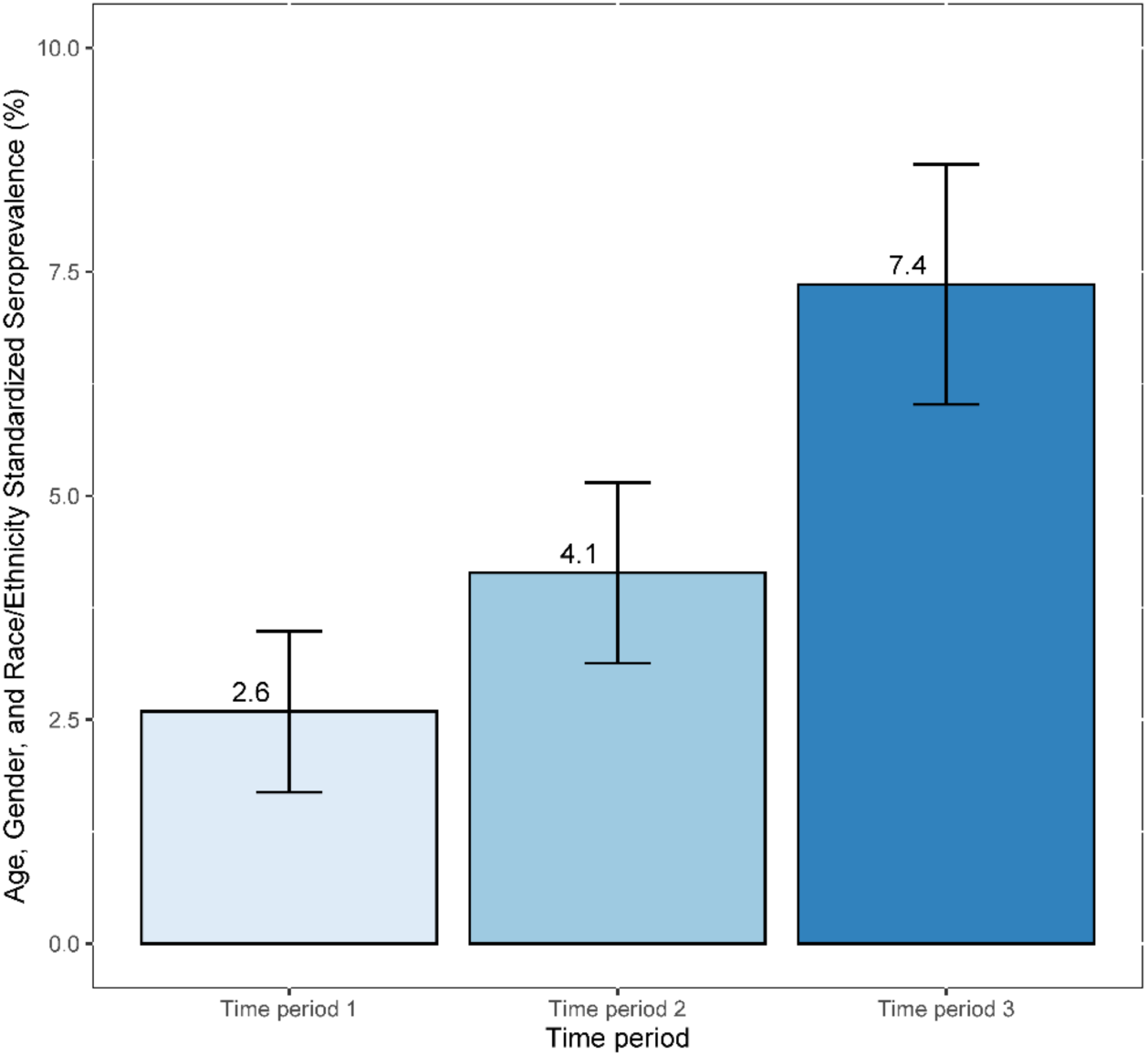
SARS-CoV-2 seropositivity rate. The age-, sex-, and race/ethnicity-adjusted seroprevalence rate are shown for each time period. Error bars indicate 95% CI.

### Comparison of racial and ethnic differences in seroprevalence by time period

Perhaps the most striking finding we observed during the course of study were differences in seroprevalence between racial and ethnic groups. During time period 1, 17.6% of Hispanic and 4.8% of Black had antibodies to CoV-2 compared to only 1.1% of white participants. During period 2, the percentage of Hispanics with CoV-2 antibodies increased to nearly 20.6% compared to 5.4% of Blacks or 2.6% of Whites. Finally, in time period 3, 23.4% of Hispanics, 8.9% of Blacks, and 5.5% of Whites had antibodies to CoV-2 (Figure 3). For each racial/ethnic group we evaluated, we observed a consistent increase in seroprevalence over the course of study; however, the difference between time periods was not significant for the Hispanic cohort due to large data variation related to relative sample sizes for this group (Table 4). During all three time periods, we noticed that Black and Hispanic populations had a higher chance of contracting the virus relative to White participants when adjusted for other variables (Table 3, Figure 4). A logistic regression fitting model showed that the increased slope of seroprevalence by time period for Hispanics was significantly lower compared to the slope for White (OR: 0.49, 95% CI: 0.3-0.78) (Table 5). In other words, although the positivity to CoV-2 was higher in Hispanic individuals compared to White individuals, this difference became smaller over the time course of the study. We obtained the same direction of the estimate for Black individuals, but there was not a statistical difference for Blacks compared to Whites (OR: 0.75, 95% CI: 0.5-1.12).

**Table 4.** Positivity of CoV-2 antibody by time period, and the temporal changing trend in the positivity overall or by the Race/Ethnicity group.

|  | Time periods 1<br>Pos. % (95% CI) | Time periods 2<br>Pos. % (95% CI) | Time periods 3<br>Pos. % (95% CI) | Time periods 2 vs. 1<br>Diff. % (95% CI) | Time periods 3 vs. 2<br>Diff. % (95% CI) | Trend test<br>*p*-value |
| --- | --- | --- | --- | --- | --- | --- |
| Overall | 3.77 (2.8, 4.95) | 4.91 (4.02, 5.92) | 8.07 (7.01, 9.23) | 1.14 (-0.24, 2.53) | 3.16 (1.73, 4.59) | <.0001 |
| Race/Ethnicity |  |  |  |  |  |  |
| White | 1.06 (0.39, 2.3) | 2.56 (1.65, 3.79) | 5.54 (4.27, 7.04) | 1.5 (0.18, 2.82) | 2.97 (1.3, 4.65) | <.0001 |
| Black | 4.84 (3.21, 6.96) | 5.42 (3.98, 7.18) | 8.87 (7.2, 10.78) | 0.58 (-1.78, 2.93) | 3.45 (1.13, 5.78) | 0.0009 |
| Hispanic | 17.57 (9.7, 28.17) | 20.63 (13.94, 28.75) | 23.36 (16.56, 31.34) | 3.07 (-8.12, 14.04) | 2.72 (-7.28, 12.73) | 0.3409 |

**Table 5.** Trend of adjusted effect of Race/Ethnicity on positivity of CoV-2 antibody.

|  | Time period increases by 1<br>OR (95% CI) | *p*-value |
| --- | --- | --- |
| Race/Ethnicity |  | 0.0314 |
| Black vs White | 0.75 (0.5, 1.12) |  |
| Hispanic vs White | 0.49 (0.3, 0.78) |  |

**Figure 3.**
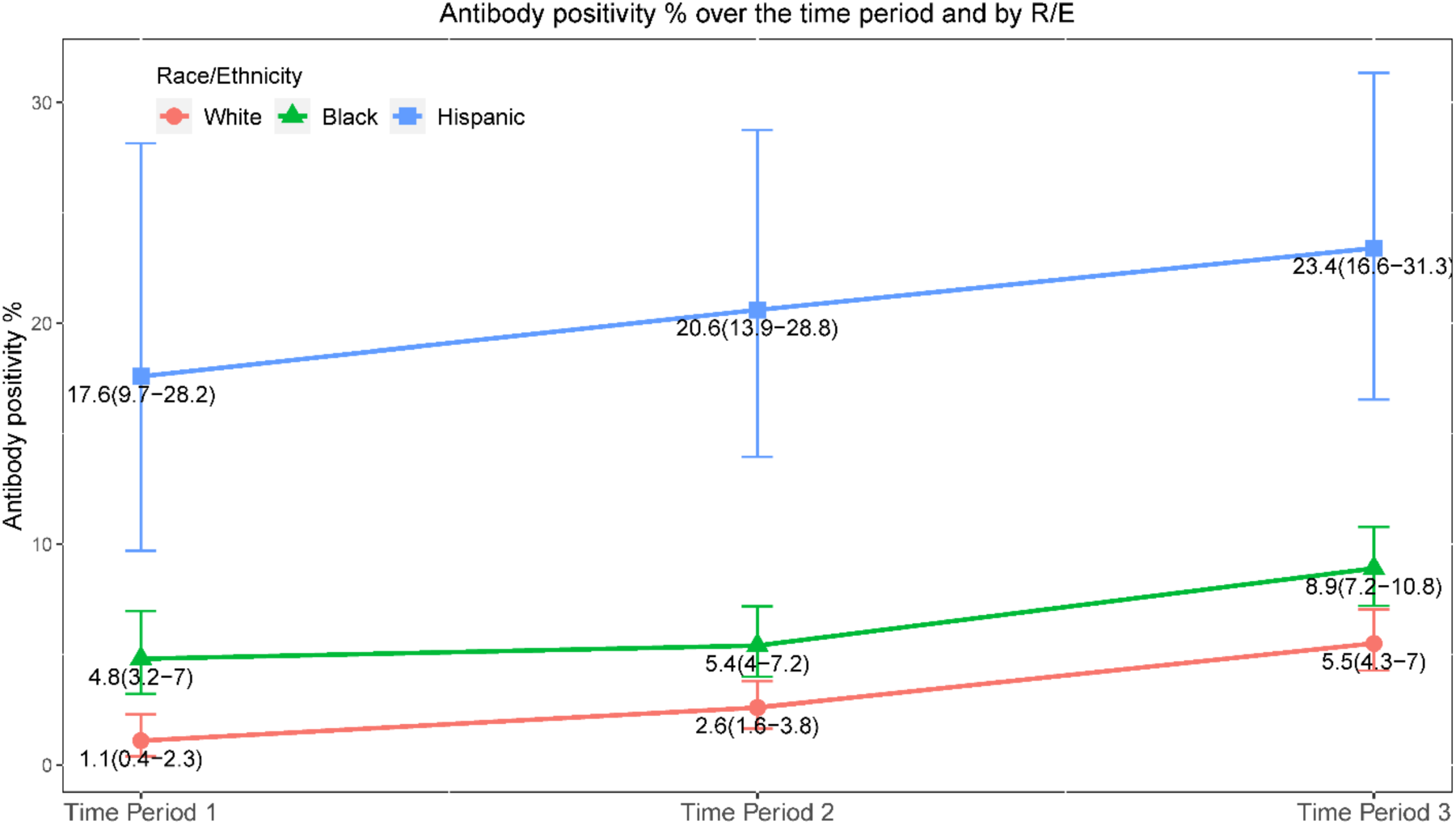
Figure 3 SARS-CoV-2 antibody positivity rates by race/ethnicity. The percentage of enrollees in different race/ethnicity groups is shown for each time period. Error bars indicate the 95% CI.

**Figure 4.**
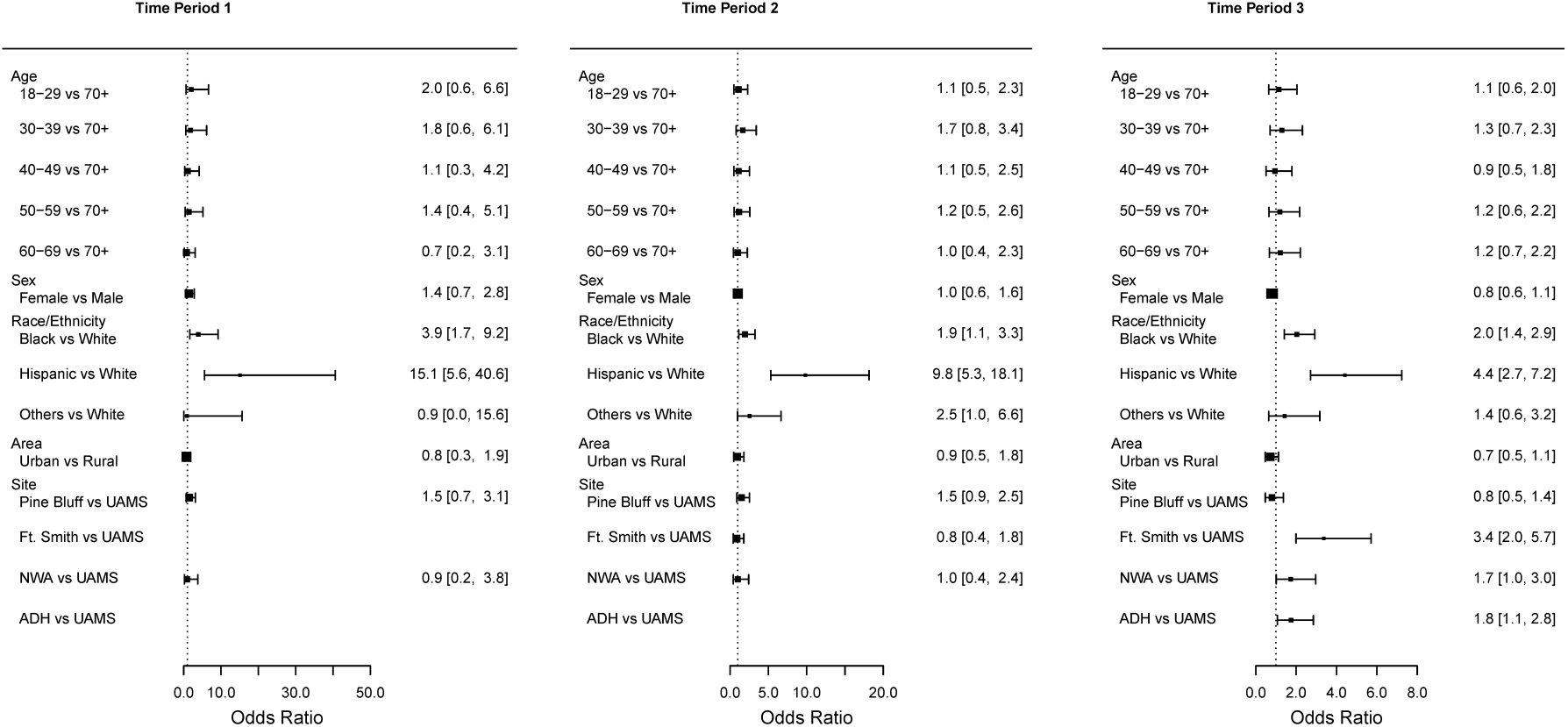
Forest plots by time period for multivariable analysis adjusted effects. Note the high Odds Ratios associated with Hispanics and Black or African Americans compared to Whites throughout the study.

### Comparison of age and gender differences in seroprevalence by time period

During the time course of this study, we noticed differences between the seroprevalence in different age groups in the univariable analysis (Table 2). During time period 1, individuals 18-29 years of age were more likely to have a positive antibody test compared to individuals greater than age 70+ (OR 3.59, 95%CI 1.12-11.52). For time period 2 and time period 3, individuals 30-39 years were more likely to have a positive antibody test compared to individuals age 70+ (Time period 2: Crude OR 2.17, 95%CI 1.08-4.36; Time period 3: Crude OR 1.85, 95% CI 1.00-3.06) (Table 2). However, with the multivariable analysis, these differences disappeared (Table 3). There were also no specific differences noted in any time periods between genders (Figure 4).

### Comparison of seroprevalence in urban and rural areas of the State

There was no statistically significant difference between seroprevalence in samples from rural and urban ZIP Code Tabulation Areas at any time point of the study. Early in the pandemic, urban areas had a slightly higher seroprevalence over rural (Time period 1: 3.9% urban, 2.9% rural; Time period 2: 5.0% urban and 4.2% rural); in time period 3, we observed a higher seroprevalence in rural over urban (7.7% urban vs. 9.7% rural). This observation is consistent with national data that urban areas may have had higher prevalence of infections in the beginning of the pandemic, but rural areas increased prevalence with time, although the effect in our study was not statistically significant (Tables 2 and 3).

### Temporal variations in CoV-2 seroprevalence

We also examined changes in seroprevalence by week. There was a gradual increase throughout the study that corresponded with increasing proportions of people infected with CoV-2 as diagnosed by molecular means. As expected, the peak of seroprevalence was noted in time period 3 towards the end of the study period with values ranging from 5.3% to 13.7% (Figure 5). Rates for racial/ethnic groups were monitored by time period and also increased accordingly with time. Seroprevalence of CoV-2 antibodies amongst Black and Hispanic individuals was consistently higher throughout the course of the study as compared to Whites. The trend test (Table 4) showed that the seroprevalence significantly increased from time period 1 to time period 3 among the White (*p* trend: <.0001) and Black population (*p* trend: 0.0009). The Hispanic population had a higher seroprevalence across three time periods (*p* trend: 0.3409). In the trend of adjusted effect for race/ethnicity (Table 5), the likelihood of antibody positivity for the Hispanic group decreased compared to White population in the later time period (OR 0.49, 95%CI 0.3-0.78). While the overall seroprevalence for our study was low compared to high population centers or high-risk groups[16-19], indicating that most Arkansans were immune naïve to the virus during the study time period, seroprevalence did increase over time in all subpopulations.

**Figure 5.**
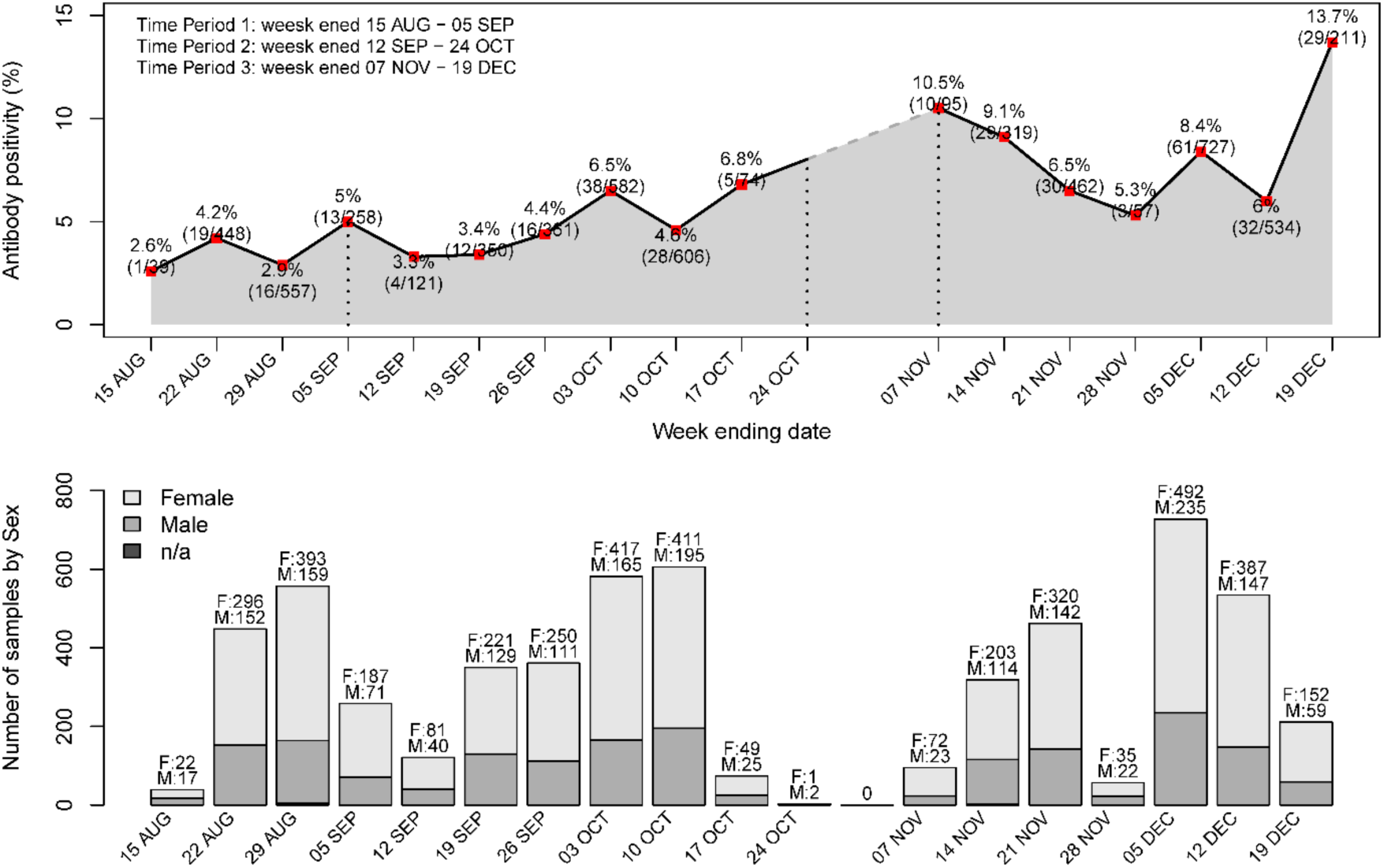
Seroprevalence in Arkansas by Week. Note the gradual increase throughout the study that corresponded with increasing proportions of people infected with CoV-2 as diagnosed by molecular means.

## DISCUSSION

Our study using medical remnant samples found that the standardized CoV-2 seroprevalence rate increased from 2.6%-7.4% in Arkansas from August to December 2020. During the last week of the study, the raw seroprevalence rate approached 14%. For comparison, the Arkansas Department of Health reported a total of 213,267 confirmed or suspected CoV-2 infections as of December 25, 2020 – roughly equivalent to 7% of the Arkansas population. Based on these numbers, it is possible that our sampling detected many asymptomatic and untested people who were previously infected with CoV-2 in our state. Many people with CoV-2 infection, including those with mild or asymptomatic infections that are likely driving the disconnect observed between the seroprevalence and acute CoV-2 infections, may go on to develop post-COVID conditions such as “long COVID” and multiorgan dysfunction. Therefore, it is imperative that seroprevalence data be coupled with acute testing results from PCR and other modalities to inform healthcare resource allocation and manage the true burden of acute and post-COVID morbidity as the pandemic progresses.

We found higher seroprevalence in Hispanics and Blacks compared to whites throughout the study. However, a temporal trend of the data showed that the increase in the white population was more than that in the Hispanics or Blacks during the same time period. Anecdotally, Hispanics and Blacks in our state were noted to have higher rates of PCR positive tests early in the course of the time periods. Ultimately, the rates of Hispanics and Blacks who had positive PCR tests leveled over the course of our study, which explains the lower odds ratios and change in effect size associated with these groups.

Our findings that younger age, race, and ethnicity correlate with a higher rate of previous CoV-2 infection in Arkansas agree with the American Red Cross data for southern states and the broader US population as a whole[21]. Any attempt to explain the racial/ethnic disparities observed in this study would be speculative, but the data fit with a broader theme highlighting the need to understand biologic, social, and demographic factors that impact health in underrepresented minority populations.

In addressing potential differences between rural and urban areas of the state, our findings suggest that CoV-2 has spread uniformly across urban and rural areas of the state. This finding was somewhat surprising given that urban areas were more greatly impacted in the northeast and northwest US, and in southern cities such as Houston and New Orleans. However, it is worth noting that Arkansas was home to a rural super-spreader event in March of 2020[23]. These data suggest that those in rural areas of the state are just as likely to have been infected with CoV-2 as those in urban population centers.

Benefits and limitations of convenience sampling techniques have been discussed elsewhere[20]. It is possible that our sampling method could favor subjects who were more ill (e.g., individuals who were hospitalized) or more willing to leave their homes (e.g., individuals when were evaluated in clinics), etc. Nonetheless, the seroprevalence observed is consistent with both reported infections and data from American Red Cross blood donations in southern US states[21] and the Centers for Disease Control Multi-State Assessment of SARS-CoV-2 Seroprevalence (MASS-C)[22], which increases confidence in our data. The American Red Cross blood study found only 2.9% seroprevalence from August to September in southern states, while the MASS-C data revealed 9.2% seroprevalence in December in Arkansas. It should be noted that these earlier studies had no[17] or low (n=1071 Arkansas samples)[22] representation from Arkansas. Taken together, the data support the conclusion that the while seroprevalence rates demonstrate that more Arkansans have been infected with CoV-2 than previously recognized, the majority of the population in a representative rural southern state was not infected by CoV-2 from March to December of 2020. This, in combination with slow vaccine uptake despite rapid distribution of vaccinations across the State, has left many people vulnerable to CoV-2 infection with variants of concern. In fact, this spring, Arkansas is experiencing an uptick in cases, ranking second in highest number of cases per 100,000 people.

## CONCLUSIONS

Seroprevalence studies play a critical in defining the scope of the CoV-2 pandemic. In a state with a large rural populace, serologic analysis of remnant samples suggest that CoV-2 infection has been more widespread than acute testing reflects. Additionally, Hispanic and Black Arkansans had disproportionately higher rates of CoV-2 infections. This study highlights the need to understand factors that impact health in underrepresented minority populations and the key role seroprevalence will play in understanding the CoV-2 pandemic.

## Data Availability

Data from this manuscript is readily available for review with the proper approvals.

## Acknowledgements

The Translational Research Institute at UAMS provided support for logistics, coordination and implementation of this project, including provision of an honest broker, secure data storage and management through REDCap (NCATS/NIH 1 UL1 RR029884).

